# Remedial angioplasty or stenting for acute basilar artery occlusion: A Post Hoc Analysis of the ATTENTION Trial

**DOI:** 10.1101/2024.10.20.24315841

**Authors:** Zhiliang Guo, Xuehan Liu, Shuhong Yu, Chunrong Tao, Pengfei Xu, Chao Zhang, Wei Hu, Guodong Xiao

**Affiliations:** Department of Neurology, The Second Affiliated Hospital of Soochow University, Suzhou 215004, China; Department of Neurology, The First Affiliated Hospital of USTC, Division of Life Sciences and Medicine, University of Science and Technology of China, Hefei, 230001, China; Department of Encephalopathy, Suzhou Integrated Traditional Chinese and Western Medicine Hospital, Suzhou 215101, China

**Keywords:** Basilar artery occlusion, endovascular treatment, balloon angioplasty, stent implantation, atherosclerosis

## Abstract

**BACKGROUND:** Whether remedial angioplasty or stenting (RAS) improves clinical outcomes of patients with basilar artery occlusion (BAO) undergoing endovascular treatment (EVT) is controversial. This study aimed to investigate the impact of RAS on prognosis in acute BAO.

**METHODS:** This post-hoc analysis derived from the ATTENTION study. Patients undergoing EVT were categorized in the RAS group if they received balloon angioplasty, stent implantation, or balloon plus stenting. The primary outcome was favorable outcome (modified Rankin Scale score of 0–3) at 90 days. Safety outcomes included death within 90 days and any intracranial hemorrhage within 24 h. Control of confounders using inverse probability processing weighting (IPTW).

**RESULTS:** A total of 221 patients with BAO experiencing EVT were included, of whom 104 (47.06%) received RAS. Multivariate analysis by IPTW adjustment showed that there was no significant difference in favorable outcome between the two groups (OR, 0.81, 95% CI, 0.55, 1.19; *P*=0.282). But RAS was associated with decreased risk of excellent outcome (OR, 0.67; 95% CI, 0.45-0.98; *P*=0.042) and functional independence (OR, 0.64; 95% CI, 0.42-0.98; *P*=0.043). Regarding safety outcomes, the RAS group had a significantly lower incidence of any intracranial hemorrhage within 24 h (OR, 0.52; 95% CI, 0.28-0.92; *P*=0.027). However, there was little difference in 90-day mortality between the two groups. In non-atherosclerotic populations, RAS is usually associated with a worse clinical prognosis; however, this phenomenon has not been found in atherosclerotic populations (all *P*<0.05 for the stroke etiology×RAS interaction). In RAS patients, we find that there was no difference in functional or safety outcomes between only balloon angioplasty (52 patients) compared with other remedy group (52 patients).

**CONCLUSIONS:** Despite the fact that RAS is secure, it didn’t further improve favorable outcomes of patients with BAO undergoing EVT; particularly in the non-atherosclerotic population. In patients with RAS, balloon angioplasty alone is an appropriate remedial technique, additional stent implantation may not be required.

## Introduction

With the publication of two high-quality clinical studies, ATTENTION and BAOCHE, there is hope and light at the endovascular treatment (EVT) of posterior circulation basilar artery occlusion (BAO).^1, 2^ Of note, both studies had high rates of acute angioplasty with balloon or stenting, with ATTENTION having 47.06% and BAOCHE having 55%.^1, 2^ The reason for this may be related to the fact that both studies were done in China and that posterior circulation stroke in the Asian population are more often combined with atherosclerosis.^3–6^ However, the evidence on the effectiveness and safety of emergency remedial angioplasty or stenting (RAS) is limited and the results vary;^5–9^ therefore, further studies are needed to clarify 1) how effective and safe RAS is in patients undergoing EVT for acute BAO; 2) is there a difference in clinical prognosis between RAS performed in atherosclerotic versus non-atherosclerotic populations; and 3) whether or not there is a difference between balloon remedy alone and other remedy modalities (stenting or balloon plus stenting) in patients undergoing RAS. If there is no difference in prognosis between the different remedies, balloon angioplasty alone is an appropriate remedial technique; additional stent implantation may not be required.

To address these questions, we performed a post hoc analysis using data from the ATTENTION clinical trial,^1^ aiming to provide information on whether or not to perform RAS in patients with acute BAO undergoing EVT, as well as the optimal RAS-suitable population and appropriate remedial techniques.

## Methods

### Data Availability

Data that reproduced the results are available to researchers from the corresponding author upon reasonable request.

### Study Design and Patient Selection

This study was a post hoc analysis of the ATTENTION trial, which was a multicenter, prospective, randomized, controlled trial of EVT for BAO at 36 centers in China from February 2021 to September 2022. Patients presenting within 12 h of symptom onset were assigned, in a 2:1 ratio, to the EVT or best medical care groups (control). This trial was registered at https://www.clinicaltrials.gov/, number NCT04751708.^1^ ATTENTION was approved by the ethics committee of all participating sites. Written informed consent from all patients or their legally authorized representative were obtained. Details of the inclusion/exclusion criteria and interventions of the ATTENTION trial have been reported.^1, 10^ For this post-hoc analysis, only patients undergoing EVT were included (Figure1: Flowchart of patients selected for analysis). This study followed the Strengthening the Reporting of Observational Studies in Epidemiology reporting guideline.^10, 11^

### RAS Definition and Classification of Stroke Cause

Patients were categorized in the RAS group if they received one of the three remedial treatments: balloon angioplasty, stent implantation, or balloon plus stenting. The choice of specific remedial timing and remedial strategies (including balloon angioplasty, stent implantation, or combinations of these approaches) were left to the discretion of the treating team. And we further categorized RAS patients into balloon angioplasty group and non-balloon angioplasty remedy group (including stenting and balloon combined stenting). Stroke etiology in the post hoc analysis was categorized into atherosclerotic group and non-atherosclerotic group. Atherosclerotic group corresponds to large-artery atherosclerosis in the TOAST criteria. The non-atherosclerosis group corresponds to the cardioembolism, undetermined cause and other determined cause.^1, 10^

### Outcomes

The primary outcome was favorable outcome, defined as a score of 0 to 3 on the modified Rankin scale (mRS) at 90 days. The secondary outcomes were excellent outcome (90-day mRS score 0–1), functional independence (90-day mRS score 0–2), the mRS shift analysis at 90 days, National Institutes of Health Stroke Scale (NIHSS) score at 24 hours, and at 72 hours, successful recanalization at final angiogram (modified Thrombolysis in Cerebral Infarction≥2b), and the number of passes.^1, 10^ Safety outcomes included intracranial hemorrhage (ICH) incidence based on the Heidelberg classification criteria (any ICH) and mortality at 90-days.^12^

### Statistical Analysis

Categorical data are expressed as percentage and frequency, and continuous variables data are reported as median and interquartile range (IQR). Inverse probability of treatment weighting (IPTW) was used to control for confounders. Standardized mean difference (SMD) were used to assess covariate balance after matching and to near or below 0.2 indicating that observed covariate imbalance was largely removed between the RAS vs No RAS. Multivariate analyses were used to explore the effect of RAS on clinical prognosis. Interaction tests of stroke etiology (atherosclerotic versus non-atherosclerotic populations)×RAS were used to assess the heterogeneity of associations between subgroups. All statistical analyses were performed using R (version 4.3.2). Two-tailed *P*<0.05 was considered statistically significant.

## Results

### Baseline Characteristics

In the ATTENTION trial, 226 patients were assigned to the EVT group; 3 patients in the EVT group crossed over to the control group, two patients in the EVT group underwent diagnostic angiography only, owing to complete recanalization after intravenous thrombolysis. Therefore, 221 patients with BAO underwent EVT were included in this post hoc analysis. There were 104 patients in the RAS group, and 117 patients in the No RAS group. Of the 104 patients in the RAS group, 52 were in the only balloon angioplasty group and 52 were in the other remedy group (5 with stenting, 47 with balloon + stenting). A flowchart of patient selection is shown in Figure 1.

**Figure 1.**
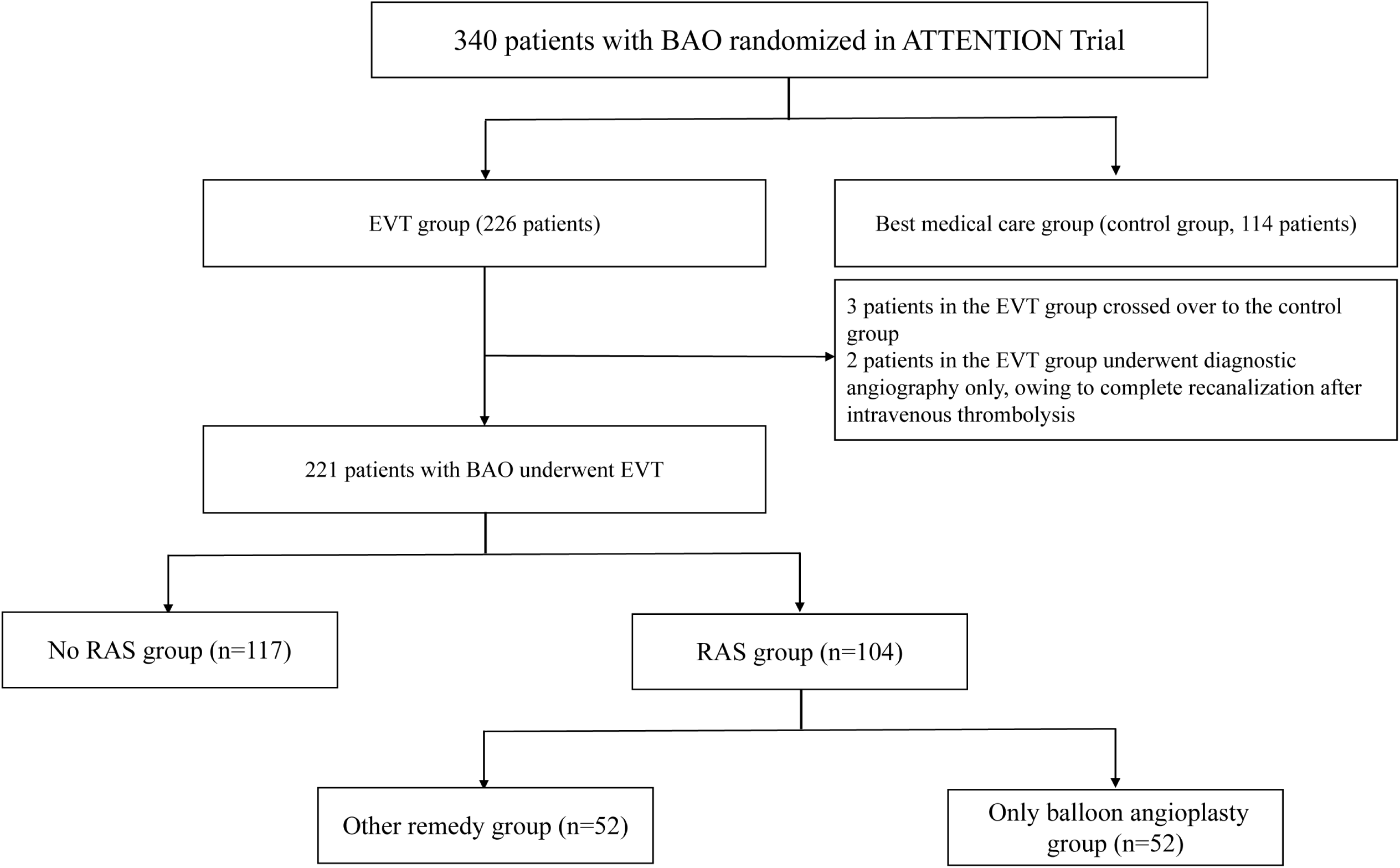
Flow diagram of patient allocation by RAS and remedial technique in the ATTENTION trial (Trial of Endovascular Treatment of Acute Basilar-Artery Occlusion). BAO indicates basilar-artery occlusion; EVT, endovascular treatment; RAS, remedial angioplasty or stenting.

The patients’ mean age was 65.9 years, 148 (67%) were men, and the median baseline NIHSS score was 24 points (IQR 15–35). As shown in Table 1, except male, mRS 1-2 at baseline, systolic blood pressure, diastolic blood pressure, intravenous thrombolysis, hypercholesterolemia, smoking, and stroke history, the differences of all covariates were statistically significant before matching by IPTW (*P*<0.05). After IPTW matching, the differences of all covariates in the RAS and No RAS groups were not statistically significant; and most of the variables had an SMD value near or below 0.2 (Figure S1), and the balance was significantly improved.

**Table 1.** Baseline characteristics of patients undergoing EVT with RAS versus no RAS.

|  | Before IPTW |  |  |  | After IPTW |  |  |  |
| --- | --- | --- | --- | --- | --- | --- | --- | --- |
| Characteristic | No RAS<br>(N=117) | RAS<br>(N=104) | *P* value | SMD | No RAS<br>(N=117) | RAS<br>(N=104) | *P* value | SMD |
| Age (y), median (p25-p75) | 69.0 [61.0, 75.0] | 65.5 [55.8, 71.0] | 0.012 | 0.297 | 67.0 [57.4, 74.0] | 66.0 [56.0, 73.0] | 0.703 | 0.043 |
| Male, n (%) | 73 (62.4) | 75 (72.1) | 0.164 | 0.208 | 70.8 (33.8) | 59.4 (28.5) | 0.525 | 0.113 |
| NIHSS score, median (p25-p75) | 30.0 [16.0, 35.0] | 20.5 [14.8, 33.2] | 0.003 | 0.414 | 25.0 [15.0, 35.0] | 22.0 [16.9, 35.0] | 0.628 | 0.094 |
| pc-ASPECTS, median (p25-p75) | 10.0 [8.0, 10.0] | 9.0 [8.0, 10.0] | 0.010 | 0.298 | 9.0 [7.0, 10.0] | 9.0 [8.0, 10.0] | 0.902 | 0.047 |
| GCS, median (p25-p75) | 7.0 [5.0, 10.0] | 10.0 [7.0, 12.0] | 0.001 | 0.423 | 8.0 [6.0, 12.0] | 9.0 [6.0, 12.0] | 0.459 | 0.101 |
| mRS 1-2 at baseline, n(%) | 13 (11.1) | 11 (10.6) | >0.999 | 0.017 | 21.2 (10.1) | 21.8 (10.5) | 0.936 | 0.012 |
| Systolic blood pressure, median (p25-p75), mmHg | 143.0 [133.0, 165.0] | 155.5 [135.0, 170.0] | 0.325 | 0.137 | 145.0 [132.0, 168.3] | 150.0 [135.5, 164.0] | 0.605 | 0.100 |
| Diastolic blood pressure, median (p25-p75), mmHg | 85.0 [76.0, 94.0] | 83.0 [77.8, 97.0] | 0.800 | 0.053 | 86.0 [76.5, 95.0] | 80.0 [77.0, 90.0] | 0.217 | 0.190 |
| Atherosclerosis, n(%) | 22 (18.8) | 84 (80.8) | <0.001 | 1.579 | 93.6 (44.6) | 105.7 (50.8) | 0.526 | 0.123 |
| Onset-to-Reperfusion, median (p25-p75), min | 6.6 [4.6, 8.7] | 7.5 [5.5, 9.3] | 0.018 | 0.269 | 6.9 [5.3, 8.5] | 7.7 [5.5, 8.8] | 0.305 | 0.075 |
| Intravenous thrombolysis, n (%) | 38 (32.5) | 29 (27.9) | 0.552 | 0.100 | 65.4 (31.2) | 57.3 (27.5) | 0.660 | 0.080 |
| Risk factors, n (%) |  |  |  |  |  |  |  |  |
| Hypertension | 76 (65.0) | 82 (78.8) | 0.033 | 0.313 | 157.3 (75.0) | 155.2 (74.6) | 0.952 | 0.011 |
| Hypercholesterolemia | 25 (21.4) | 33 (31.7) | 0.111 | 0.236 | 47.0 (22.4) | 62.5 (30.0) | 0.369 | 0.174 |
| Diabetes mellitus | 22 (18.8) | 26 (25.0) | 0.341 | 0.150 | 31.1 (14.8) | 52.2 (25.1) | 0.155 | 0.26 |
| Atrial fibrillation | 31 (26.5) | 2 (1.9) | <0.001 | 0.752 | 32.8 (15.7) | 19.6 (9.4) | 0.510 | 0.189 |
| Smoking | 28 (23.9) | 33 (31.7) | 0.253 | 0.175 | 61.3 (29.2) | 48.8 (23.4) | 0.437 | 0.132 |
| Coronary artery disease | 29 (24.8) | 6 (5.8) | <0.001 | 0.548 | 34.9 (16.7) | 27.2 (13.1) | 0.676 | 0.101 |
| Stroke history | 28 (23.9) | 26 (25.0) | 0.978 | 0.025 | 62.8 (29.9) | 61.7 (29.7) | 0.977 | 0.006 |
| Occlusion site, n (%) |  |  |  |  |  |  |  |  |
| Proximal basilar artery | 26 (22.2) | 42 (40.8) | 0.001 | 0.557 | 52.3 (24.9) | 82.9 (40.1) | 0.107 | 0.494 |
| Middle basilar artery | 31 (26.5) | 29 (28.2) |  |  | 50.6 (24.1) | 61.8 (29.8) |  |  |
| Distal basilar artery | 51 (43.6) | 21 (20.4) |  |  | 78.5 (37.4) | 36.0 (17.4) |  |  |
| Vertebral artery V4 | 9 (7.7) | 11 (10.7) |  |  | 28.4 (13.5) | 26.3 (12.7) |  |  |
| Stroke cause, n (%) |  |  |  |  |  |  |  |  |
| Atherosclerosis | 22 (18.8) | 84 (80.8) | <0.001 | 1.641 | 93.6 (44.6) | 105.7 (50.8) | 0.518 | 0.246 |
| Cardioembolism | 43 (36.8) | 3 (2.9) |  |  | 45.9 (21.9) | 26.4 (12.7) |  |  |
| Other or unknown etiology | 52 (44.4) | 17 (16.3) |  |  | 70.2 (33.5) | 76.0 (36.5) |  |  |

EVT, endovascular treatment; GCS, Glasgow coma scale; IQR, interquartile range; IPTW, inverse probability processing weighting; mRS, modified Rankin Scale; NIHSS, National Institutes of Health Stroke Scale; pc-ASPECTS, posterior circulation Acute Stroke Prognosis Early Computed Tomography Score; RAS, remedial angioplasty or stenting.

### Outcomes of patients undergoing EVT with RAS versus No RAS

The favorable outcome was 46.2% in the RAS group; 45.3% in the No RAS group. There were also no statistical differences between the two groups in other outcome indicators, such as excellent outcome, functional independence, mRS, NIHSS score at 24 hours, and at 72 hours, successful recanalization, the number of passes, any ICH and mortality, when unadjusted (all *P*>0.05; Table 2). Multivariate analysis showed that RAS was not an independent risk factor for favorable outcome after IPTW (OR, 0.81, 95% CI, 0.55, 1.19; *P*=0.282]; but was associated with decreased risk of excellent outcome (OR, 0.67; 95% CI, 0.45-0.98; *P*=0.042) and functional independence (OR, 0.64; 95% CI, 0.42-0.98; *P*=0.043). Patients in the RAS group had higher recanalization rates (OR, 3.20; 95% CI, 1.55-7.16; *P*=0.003) and a greater number of passes (OR, 1.41; 95% CI, 1.13-1.76; *P*=0.002). Regarding safety outcomes, the RAS group had a significantly lower incidence of any intracranial hemorrhage within 24 h (OR, 0.52; 95% CI, 0.28-0.92; *P*=0.027). However, there was little difference in 90-day mortality between the two groups (Table 2).

**Table 2.** Outcome measures of patients undergoing EVT with RAS versus No RAS.

| Outcome variables | No RAS<br>(n = 117) | RAS<br>(n = 104) | Unadjusted model |  | Adjusted model by IPTW |  |
| --- | --- | --- | --- | --- | --- | --- |
|  |  |  | Effect size (95%<br>CI) | P value | Effect size (95% CI) | P value |
| Primary outcome<sup>a</sup> |  |  |  |  |  |  |
| mRS 0-3 at 90d, n(%) | 53 (45.3) | 48 (46.2) | 1.04(0.61,1.76) | 0.899 | 0.81(0.55,1.19) | 0.282 |
| Secondary outcomes |  |  |  |  |  |  |
| mRS 0-1 at 90d, n(%) | 60 (51.3) | 52 (50.0) | 0.95(0.56,1.61) | 0.849 | 0.67(0.45,0.98) | 0.042 |
| mRS 0-2 at 90d, n(%) | 37 (31.6) | 35 (33.7) | 1.1(0.62,1.93) | 0.748 | 0.64(0.42,0.98) | 0.043 |
| mRS 0-3 at 90d, n(%) | 53 (45.3) | 48 (46.2) | 1.04(0.61,1.76) | 0.899 | 0.81(0.55,1.19) | 0.282 |
| mRS at 90 d, median (IQR)<sup>a</sup> | 4.0 [2.0, 6.0] | 4.5 [2.0, 6.0] | 1.00(0.86,1.15) | 0.973 | 1.10(0.96,1.25) | 0.164 |
| NIHSS score at 24 h, median (IQR)<sup>a</sup> | 23.0 [8.0, 35.0] | 19.0 [7.0, 36.0] | 0.96(0.81,1.14) | 0.612 | 0.90(0.76,1.07) | 0.235 |
| NIHSS score at 72h, median (IQR)<sup>a</sup> | 17.0 [4.0, 35.0] | 15.0 [4.0, 37.0] | 0.98(0.79,1.22) | 0.857 | 0.90(0.73,1.13) | 0.373 |
| Successful recanalization at final angiogram (mTICI≥2b), n(%) | 108 (92.3) | 98 (94.2) | 1.36(0.47,4.19) | 0.572 | 3.20(1.55,7.16) | 0.003 |
| Number of passes<sup>a</sup> | 1.0 [1.0, 2.0] | 1.0 [1.0, 2.2] | 1.20(0.99,1.45) | 0.060 | 1.41(1.13,1.76) | 0.002 |
| Safety outcomes |  |  |  |  |  |  |
| Death within 90d, n(%) | 43 (36.8) | 39 (37.5) | 1.03(0.6,1.78) | 0.909 | 1.44(0.98,2.12) | 0.066 |
| Any ICH within 24 h, n(%) | 17 (14.5) | 14 (13.5) | 0.92(0.42,1.96) | 0.819 | 0.52(0.28,0.92) | 0.027 |

EVT, endovascular treatment; ICH, intracranial hemorrhage; IQR, interquartile range; IPTW, inverse probability processing weighting; mRS, modified Rankin Scale; NIHSS, National Institutes of Health Stroke Scale; RAS, remedial angioplasty or stenting.
<sup>a</sup>The effect size was calculated using a generalized linear model (link function is quasipoisson) and performing an exponential transformation.

**Table 3.** Outcomes of patients undergoing EVT with balloon versus others in RAS.

| Outcome variables | Ball<br>(n = 52) | Other<br>(n = 52) | Unadjusted model |  | Adjusted model |  |
| --- | --- | --- | --- | --- | --- | --- |
|  |  |  | Effect size (95%<br>CI) | P value | Effect size (95% CI) | P value |
| Primary outcome |  |  |  |  |  |  |
| mRS 0-3 at 90d, n(%) | 26 (50.0) | 22 (42.3) | 0.67(0.28,1.6) | 0.373 | 0.73(0.34,1.59) | 0.432 |
| Secondary outcomes |  |  |  |  |  |  |
| mRS 0-1 at 90d, n(%) | 29 (55.8) | 23 (44.2) | 0.59(0.25,1.39) | 0.231 | 0.63(0.29,1.36) | 0.24 |
| mRS 0-2 at 90d, n(%) | 16 (30.8) | 19 (36.5) | 1.42(0.54,3.84) | 0.477 | 1.3(0.57,2.95) | 0.534 |
| mRS at 90 d, median (IQR)<sup>a</sup> | 3.5 [2.0, 6.0] | 5.0 [1.0, 6.0] | 0.98(0.79,1.21) | 0.832 | 1.01(0.81,1.24) | 0.963 |
| NIHSS score at 24 h, median (IQR)<sup>a</sup> | 19.0 [7.0, 35.0] | 22.0 [7.8, 37.0] | 0.97(0.75,1.24) | 0.796 | 1.06(0.82,1.37) | 0.673 |
| NIHSS score at 72h, median (IQR)<sup>a</sup> | 14.0 [5.5, 28.0] | 17.0 [3.0, 38.0] | 1.04(0.76,1.43) | 0.799 | 1.16(0.84,1.59) | 0.371 |
| Successful recanalization at final angiogram (mTICI≥2b), n(%) | 48 (92.3) | 50 (96.2) | 4.78(0.69,54.37) | 0.142 | 2.08(0.39,15.53) | 0.409 |
| Number of passes<sup>a</sup> | 2.0 [1.0, 2.2] | 1.0 [1.0, 2.2] | 0.81(0.62,1.06) | 0.124 | 0.85(0.65,1.12) | 0.265 |
| Safety outcomes |  |  |  |  |  |  |
| Death within 90d, n(%) | 17 (32.7) | 22 (42.3) | 1.49(0.62,3.65) | 0.378 | 1.51(0.68,3.39) | 0.312 |
| Any ICH within 24 h, n(%) | 6 (11.5) | 8 (15.4) | 1.09(0.32,3.79) | 0.888 | 1.39(0.45,4.54) | 0.567 |

EVT, endovascular treatment; ICH, intracranial hemorrhage; IQR, interquartile range; mRS, modified Rankin Scale; NIHSS, National Institutes of Health Stroke Scale; RAS,
remedial angioplasty or stenting.
<sup>a</sup>The effect size was calculated using a generalized linear model (link function is quasipoisson) and performing an exponential transformation.
Adjusted model were adjusted for age, the modified Rankin scale score before stroke onset, the time from onset to reperfusion, and NIHSS score in baseline.

### Outcomes of patients undergoing EVT with RAS versus No RAS by stroke etiology

In non-atherosclerotic populations, RAS is usually associated with a worse clinical prognosis (favorable outcome: OR, 0.44; 95%CI, 0.25-0.77; *P*=0.005. excellent outcome: OR, 0.38; 95%CI, 0.22-0.67; *P*<0.001. functional independence: OR, 0.22; 95%CI, 0.10-0.45; *P*<0.001. mRS: effect size, 1.30; 95%CI, 1.10-1.54; *P*=0.003); however, this phenomenon has not been found in atherosclerotic populations (*P* values for the stroke etiology×RAS interaction were 0.003, 0.010, <0.001, and 0.007 for favorable outcome, excellent outcome, functional dependence, and mRS, respectively).

In the non-atherosclerotic populations, higher mortality within 90 days was observed in patients treated with RAS compared with No RAS (60.0% versus 35.8%), with *P* <0.001 for interaction analysis (Figure 2). The rate of any ICH in non-atherosclerotic patients with RAS, and No RAS was 5%, and 13.7%, respectively, and *P* values for the stroke etiology×RAS interaction was 0.043 (Figure 2).

**Figure 2.**
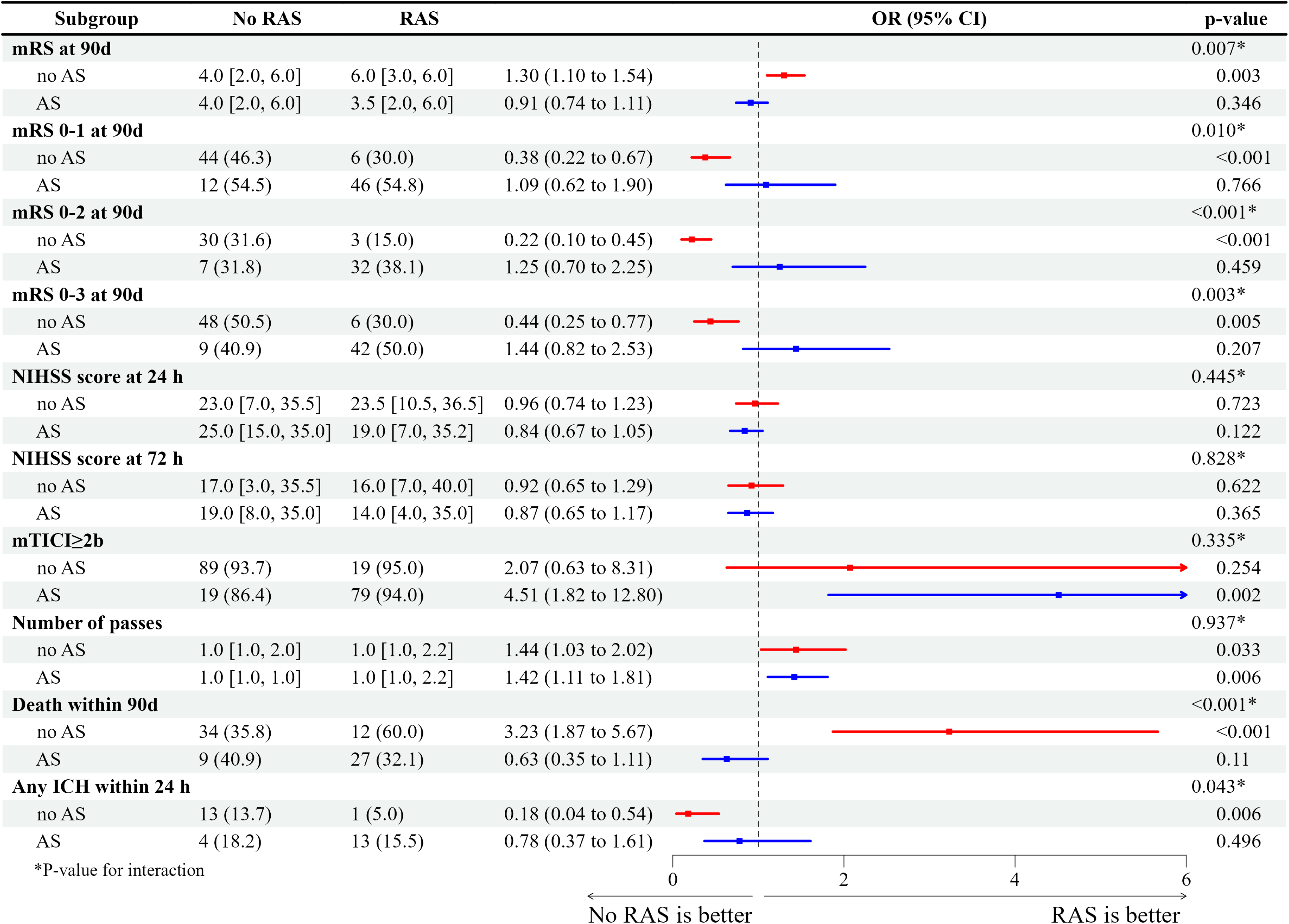
Outcomes of patients undergoing EVT with RAS versus No RAS by stroke etiology. AS indicates atherosclerotic populations; EVT, endovascular treatment; ICH, intracranial hemorrhage; mRS, modified Rankin Scale; mTICI, modified Treatment in Cerebral Infarction; NIHSS, National Institutes of Health Stroke Scale; OR, odd ratio; RAS, remedial angioplasty or stenting.

### Outcomes of patients undergoing EVT with balloon versus others in RAS

We divided the RAS patients into the only balloon angioplasty group (52 patients) and the other remedy group (52 patients). The baseline characteristics were balanced between balloon group and others group (Table S1), and we did not do further IPTW calibration. Multivariate analysis showed that there was no difference in primary outcome, secondary outcomes, or safety outcomes between only balloon angioplasty and other remedial treatments after adjusting for age, the mRS before stroke onset, the time from onset to reperfusion, and NIHSS score in baseline (all *P*>0.05).

## DISCUSSION

The purpose of this post hoc analysis of the ATTENTION clinical trial was to assess the impact of RAS on the clinical prognosis of acute BAO patients treated with EVT. The results of this article show that: 1) RAS was not associated with a further improved proportion of favorable outcome (mRS 0-3) and was associated with a reduced proportion of excellent outcome (mRS 0-2) and functional independence (mRS 0-2) in BAO patients treated with EVT. Regarding safety outcomes, RAS was not independently correlated with 90-day mortality, although it was associated with lower any ICH. 2) In the non-atherosclerotic population, RAS was associated with poorer functional prognosis, either favorable outcome, mRS, excellent outcome, or functional independence. *P* values for the stroke etiology*RAS interaction test were less than 0.05. 3) Further, we divided the RAS population into only balloon angioplasty subgroup and the other remedy group (balloon + stenting or stenting), and found no difference in functional prognosis or safety outcomes (90-day mortality and any ICH) between the two groups. Suggesting that if angioplasty remedial treatment is to be performed, balloon angioplasty alone may be an appropriate choice. In summary, atherosclerotic populations and balloon angioplasty alone may be appropriate remedial populations and appropriate remedial techniques, respectively. This also needs to be confirmed by studies with larger samples.

Because of the prevalence of intracranial atherosclerotic stenosis in the Asian population, the rate of reocclusion after EVT is high, ranging from 36.7% to 77.3%.^13–15^ About 30% of Chinese patients with acute stroke due to intracranial large artery occlusion have unsuccessful EVT, which may also require intracranial angioplasty as a remedial measure.^16^ The results of several studies have demonstrated that remedial stenting or balloon dilatation in patients with acute large vessel occlusion who have failed EVT (modified Rankin Scale <2b) improves the rate of good prognosis at 3 months and does not increase the incidence of symptomatic ICH or the rate of morbidity and mortality.^6–9^ However, there is a lack of high-quality research evidence, especially in the posterior circulation. We first analyzed the effect of RAS on the prognosis of acute BAO patients treated with EVT, and our results were partially consistent with previously published results in that RAS did not improve the primary end point metrics or increase safety adverse events.^5–9^

Although there are other rare causes, such as arterial dissection, that lead to the need for remediation after EVT,^17, 18^ the majority of reasons for patient remediation remain residual stenosis due to atherosclerosis.^5–9^ We therefore divided our patients into an atherosclerotic and a non-atherosclerotic population and found that for the non-atherosclerotic population, RAS was associated with a worse clinical prognosis, both in terms of the primary endpoint and the secondary endpoint. However, in the atherosclerotic population, there was a trend for RAS to improve the good prognosis of patients. These results suggest that the atherosclerotic but not non-atherosclerotic population may be a suitable population for remediation, this conclusion depends on larger samples in the future to confirm our conjecture.

Commonly utilized remedies for RAS include balloon dilatation alone, stent implantation alone, and balloon dilatation plus stent implantation.^1, 5, 16^ Previous studies on elective non emergency surgery for intracranial stenosis have found that intracranial stenting did not further prevent stroke events and improve patient prognosis.^19–21^ Recently, balloon angioplasty (not stenting) plus aggressive medical management significantly lowered the risk of a composite outcome of any stroke or death within 30 days or an ischemic stroke or revascularization of the qualifying artery after 30 days through 12months.^22^ The above results suggest that simple balloon dilatation may be an appropriate intracranial management option. As for addressing this issue in terms of emergency surgery, the ANGEL-REBOOT research discovered that remedial treatment did not further improve patient prognosis when the various remedies were examined as a whole.^5^ We very much look forward to ANGEL-REBOOT’s subgroup analyses targeting between different remedies to assess whether the specific surgical modality of remedy has an impact on prognosis. From our analysis of the posterior circulation data, balloon dilatation alone achieved similar results to other remedies, both in terms of functional and safety prognosis. This suggests that if a patient requires remedial therapy, balloon remediation alone may be sufficient, which would further save patient costs, avoid stent implantation, and simplify subsequent perioperative management and secondary prevention.

There are some limitations to this study. Firstly this was a post hoc retrospective analysis of a clinical trial, patients were not randomized according to whether they were RAS or not, and mismatched baseline information may have caused some interference in the results. For this reason, we used IPTW to calibrate confounders and performed multifactor adjusted analyses. Second, we only analyzed patients who experienced EVT post hoc, excluding those who did not. The overall sample size was small, which may have weakened our statistical power. Moreover, we analyzed an Asian population, which itself has a relatively high prevalence of atherosclerosis.^3–6, 10^ Therefore, extrapolation of our findings to other populations may be warranted with caution. Finally, this analysis was based on data from the database of the randomized controlled trial (RCT) study, and it was left to the discretion of the surgical team to decide on the specific timing of remediation, choice of remediation regimen, intraoperative venous or arterial use of GPIIb/IIIa receptor antagonists (tirofiban), and postoperative antithrombotic program. We look forward to future RCTs in the posterior circulation, a population with a higher prevalence of atherosclerotic etiologies, to examine the preferred remedial regimen and the impact of antithrombotic medications.

## CONCLUSIONS

The implementation of RAS in BAO patients treated with EVT did not further improve patient prognosis, especially in the non-atherosclerotic population, and it even exacerbated the risk of poor patient prognosis. However, there was a trend toward improved prognosis in the atherosclerotic population. The use of balloon angioplasty alone achieved similar results to other remedies. And it is expected that future RCTs will further explore the appropriate population and technique for remedial therapy.

## Acknowledgments

The authors are grateful to all patients and investigators in the ATTENTION trial (Trial of Endovascular Treatment of Acute Basilar-Artery Occlusion).

## Supplemental Material

Tables S1; Figure S1.

STROBE Checklist.

